## Supplementary material for "Collecting mortality data via mobile phone surveys: a non-inferiority randomized trial in Malawi": Consent script (treatment group)

| I work for IPOR in Zomba. Researchers at New York University-Abu Dhabi, Chancelor College and the London School of Hygiene and Tropical Medicine are also collaborating in the study. We are conducting a study on the impact of COVID-19 in Malawi. We are calling approximately 9000 Malawians to ask them to participate in a short interview and your telephone number has been chosen by chance.  If you agree, I would like to ask you a few questions. Your participation in this study is voluntary. From this point on, the total duration of the interview is expected to be 10-15 minutes. Your answers might help us better understand how COVID-19 has affected the lives of Malawians.  The interview will consist of questions about yourself and about your relatives (for example, the members of your household or your parents). Some of these questions will focus on deaths that may have occurred amongst these relatives in the recent past.  You can choose not to answer any of the questions that I will ask you. You are free to pause or interrupt the interview at any time. Note also that discussing deaths amongst relatives may at times cause negative feelings. If that happens, please do let me know and we can pause the interview. I can also call you back at a later time or refer you to a trained counsellor. If you complete the interview, we will transfer 1200 Kwacha in mobile phone credit as an appreciation for your time.  All the information that we will collect during this interview may be shared with other researchers, but only after we delete your name and telephone number so that nobody can link the information to you personally.  At the end of the interview, I will give you the contact details of the person who is responsible for this study. For any further inquiries, you can also contact the research ethics committee of the University of Malawi, at (265) 0524 222.  Do you have any questions for me at this point? | | | |
| --- | --- | --- | --- |
| C1 | Would you like to participate in this study? | YES  NO | I0 |
| C2 | Note to interviewer: indicate refuser’s reaction upon declining to consent to participate. Did the person state any reason for not consenting?  INSTRUCTION: DO NOT ASK THE PERSON WHO REFUSED WHY HE/SHE REFUSED, ONLY RECORD HIS/HER REACTION  *Multiple answers are allowed.* | No reason/just hung up  No Time  No interest  Did not understand the purpose of the study  Did not want to discuss recent deaths among my relatives  Other (specify) |  |
| Closing statement: thank you again for hearing about this study. I wish you a very pleasant day. | | | |
